# Monitoring COVID-19 spread in Prague local neighborhoods based on the presence of SARS-CoV-2 RNA in wastewater collected throughout the sewer network

**DOI:** 10.1101/2021.07.28.21261272

**Authors:** Kamila Zdenkova, Jana Bartackova, Eliska Cermakova, Katerina Demnerova, Alzbeta Dostalkova, Vaclav Janda, Zuzana Novakova, Michaela Rumlova, Jana Rihova Ambrozova, Klara Skodakova, Iva Swierczkova, Petr Sykora, Dana Vejmelkova, Jiri Wanner, Jan Bartacek

## Abstract

Many reports have documented that the presence of SARS-CoV-2 RNA in the influents of municipal wastewater treatment plants (WWTP) correlates with the actual epidemic situation in a given city. However, few data have been reported thus far on measurements upstream of WWTPs, i.e. throughout the sewer network. In this study, the monitoring of the presence of SARS-CoV-2 RNA in Prague wastewater was carried out at selected locations of the Prague sewer network from August 2020 through May 2021. Various locations such as residential areas of various sizes, hospitals, city center areas, student dormitories, transportation hubs (airport, bus terminal), and commercial areas were monitored together with four of the main Prague sewers. The presence of SARS-CoV-2 RNA was determined by reverse transcription – multiplex quantitative polymerase chain reaction (RT-mqPCR) after the precipitation of nucleic acids with PEG8000 and RNA isolation with TRIzol™ Reagent. The number of copies of the gene encoding SARS-CoV-2 nucleocapsid (N1) per liter of wastewater was compared with the number of officially registered COVID-19 cases in Prague. Although the data obtained by sampling wastewater from the major Prague sewers were more consistent than those obtained from the small sewers, the correlation between wastewater-based and clinical-testing data was also good for the residential areas with more than 1 000 registered inhabitants. It was shown that monitoring SARS-CoV-2 RNA in wastewater sampled from small sewers could identify isolated occurrences of COVID-19-positive cases in local neighborhoods. This can be very valuable while tracking COVID-19 hotspots within large cities.

**Highlights:**

- SARS-CoV-2 RNA presence was measured at 24 locations in the Prague sewer network
- Residential areas (100–13 000 inhab.), transport hubs, hospitals etc. were included
- Consistent wastewater monitoring by RT-mqPCR took place from August 2020 – May 2021
- The sampling of major Prague sewers correlated well with clinical-based data
- Grab samples can identify COVID-19 hotspots in local neighborhoods

**Graphical abstract:** 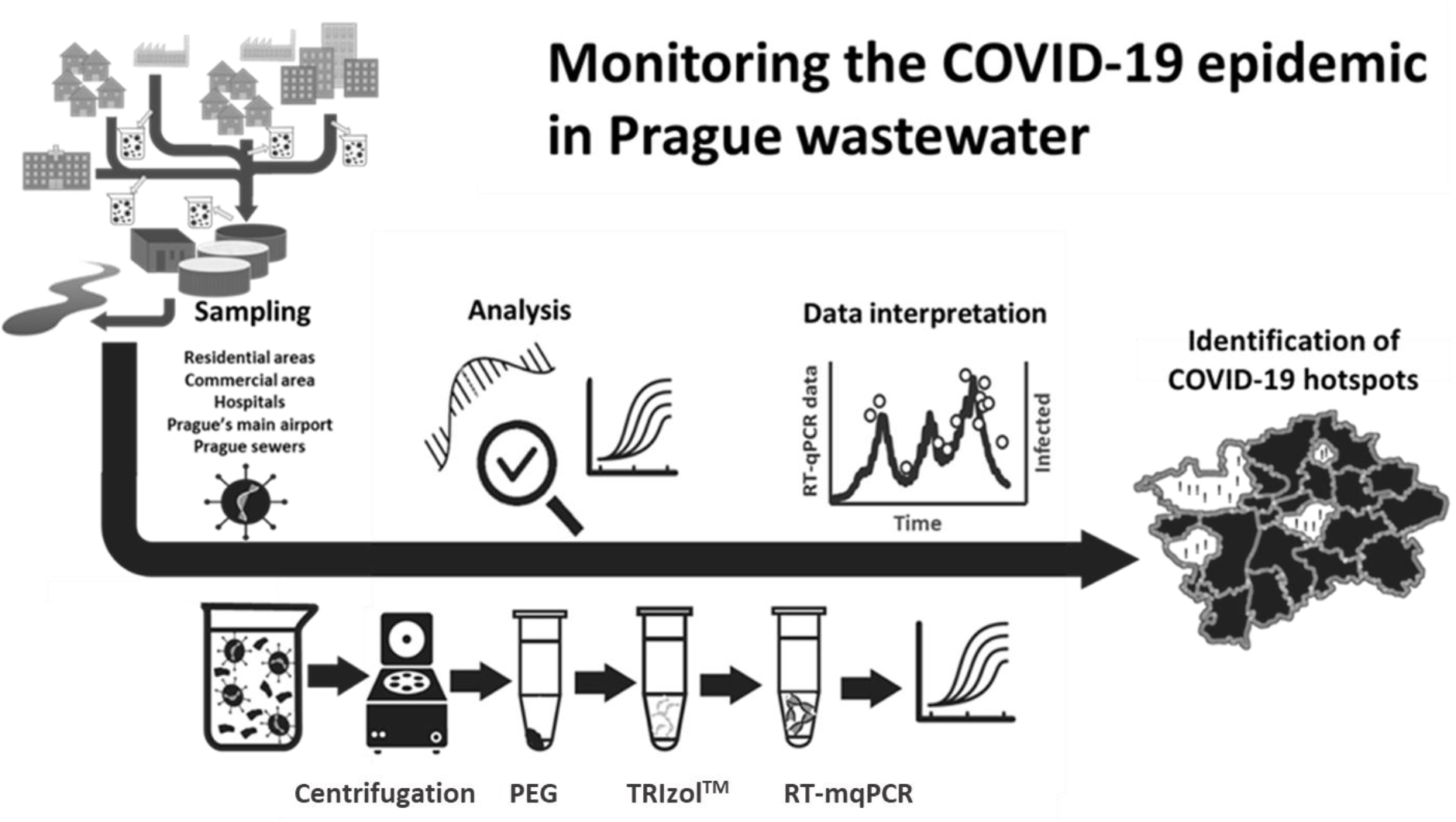

## Introduction

Even though the first wave of the COVID-19 pandemic hit Czechia mildly from March to May 2020, the numbers of patients rose sharply later in October. Since then, the Czech Republic remained one of the most severely affected countries worldwide until April 2021. At the same time, the Czech population was relatively reluctant to undergo clinical testing, resulting in an extremely high rate of positively tested patients, 30 to 40% of which were proved to be COVID-positive. This situation led to a health crisis which was extremely difficult to manage by the state authorities. In such a situation, an accurate and timely system for the monitoring of the pandemic is critically needed.

Since the first papers by Medema et al. (2020) reporting the possibility of detecting the presence of SARS-CoV-2 (the virus responsible for the COVID-19 disease) in sewage, hundreds of papers describing the correlation between viral RNA in wastewater and the COVID-19 pandemic have been published. It has been repeatedly shown that there is a close relation between gene copy numbers (typically genes E, N1, N2, N3, S, or RdRp) in wastewater and the actual epidemic situation in the respective catchment area (Heijnen et al. 2021, Randazzo et al. 2020, Saththasivam et al. 2021). Moreover, this relation can be mathematically modeled (Hart and Halden 2020).

Indeed, SARS-CoV-2 RNA fragments (genes N1, N2, and N3) have been detected in wastewater, at a level corresponding to a little over 25 positive cases per 1000 persons (Hong et al. 2021). This can be explained by the high production of viral particles in the feces of infected patients, which can be anywhere between 10^3^ and 10^7^ SARS-CoV-2 RNA copies in a gram of feces (Wölfel et al. 2020). Most authors estimated that around 50% of COVID-19 patients excrete SARS-CoV-2 RNA in feces (Chan et al. 2020, Cheung et al. 2020, Wong et al. 2020), but some studies suggest that this may be true for up to 100% of COVID-19 patients (Papoutsis et al. 2021).

The vast majority of SARS-CoV-2 monitoring studies in major cities all around the world focused on sewage sampling at the entrance of the cities’ wastewater treatment plants – WWTP (Ahmed et al. 2021b, Zhou et al. 2021). Sampling WWTP influent has many advantages: the sampling can be thoroughly controlled, it is easy to collect 24-hour composite samples that accurately represent sewage composition, and a large number of people are taken into account with a relatively limited number of samples. The downside of this approach is that the relation between the COVID-19 pandemic and wastewater-based data is largely averaged over the entire society while having relatively limited informative value for epidemiologists. I.e. epidemic outbreak spots within a city cannot be identified, and focused control measures cannot be taken.

If sampled upstream in the sewer network, specific neighborhoods can be targeted based on their importance for pandemic monitoring. Unfortunately, limited data have been published in this respect so far. Goncalves et al. (2021) had been detecting SARS-CoV-2 RNA in the hospital area over a short period of two weeks. Similarly Acosta et al. (2021) monitored wastewater from three hospitals showing that viral burden correlated with increasing hospitalized cases as well as hospital-associated transmissions. Ahmed et al. (2021a) sampled four main sewers in Bangladesh, but again, few samples were analyzed (6 sampling days within one month) to assess their epidemiological value. Saguti et al. (2021) reported results from 4 sampling sites throughout the city of Gothenburg (Sweden), but the sampling in these subareas only covered 4 weeks, and the correlation with epidemic data was unclear. Also, Baldovin et al. (2021) reported data for 4 sampling sites (pumping stations) throughout Padova (Italy) that each represented a large portion of the city.

Sofar the most detailed study using decentralized wastewater monitoring published very recently Mota et al. (2021) who monitored 17 locations within the sewer network of Belo Horizonte (Brazil) from May through August 2020 and showed that hotspots could be identified in the city based on data generated by decentralized sewage monitoring, rather than based on clinical data. The locations monitored in the latter study represented still relatively large catchment areas representing from approximately 12 thousand to 1.4 million inhabitants with the majority of the areas representing several tens of thousands of inhabitants. Long-term data of similar nature, preferably from even smaller locations, are very much needed from other places to confirm the usefulness of such monitoring.

From July 2020 through March 2021, we monitored the presence of SARS-CoV-2 RNA in sewage sampled biweekly at 14 locations throughout the Prague sewer network. Within these locations were residential areas with apartment buildings and family houses, commercial area, areas dominated by student dormitories, or Prague’s main airport. From late March through the end of May 2021, we expanded the monitoring to 24 locations including four of Prague’s main sewers and taking samples up to three times a week. In this paper, we report the presence of SARS-CoV-2 RNA in sewage and its correlation with the epidemic situation in Prague.

Following the preliminary study performed in summer 2020 by Mlejnkova et al. (2020) at WWTPs throughout the Czech Republic, this is the first consistent study on the presence of SARS-CoV-2 RNA in Prague wastewater. After the very recent paper by Mota et al. (2021), this is the first European study thoroughly reporting long-term data for local neighborhoods of different types within one city.

## Materials and Methods

### Sampling campaigns

The sampling took place in two main periods: (1) a long-term monitoring, where grab samples were taken every other week at 14 locations (Locations 1 to 14, Tab. 1) within the Prague sewer network and (2) an intensive monitoring (2 to 3 samples per week) in spring 2021 was done in the same locations as the long-term monitoring. In addition, the intensive monitoring included another eight local neighborhoods and four of Prague main sewers (24-hour composite samples).

**Tab.1.** Sampling sites monitored in this study.

| Location no. | Type of neighborhood | Number of officially registered residents | Type of samples | Separate sewer for stormwater |
| --- | --- | --- | --- | --- |
| **Local neighborhoods sampled within Prague's sewer system** |  |  |  |  |
| **- Location sampled from August 2020 till May 2021** |  |  |  |  |
| **1** | Family houses 1 | 105 | grab | Yes |
| **2** | Family houses 2 | 974 | grab | Yes |
| **3** | Family houses 3 | 10 152 | grab | No |
| **4** | Apartment buildings 1 | 5 869 | grab | No |
| **5** | Apartment buildings 2 | 7 075 | grab | Yes |
| **6** | Apartment buildings 3 | 1 153 | grab | Yes |
| **7** | Hospital & Residential area | 13 148 | grab | No |
| **8** | City center | 1 628 | grab | No |
| **9** | Office buildings | 99 | grab | No |
| **10** | Shopping mall | 1 478 | grab | Yes |
| **11** | University dormitories | 98 | grab | No |
| **12** | Industrial area | 2 340 | grab | No |
| **13** | Airport | n.a.* | grab/24-hours composite | Yes |
| **14** | Airport | n.a.* | grab | Yes |
| **- Locations sampled 6 weeks from April through May 2021** |  |  |  |  |
| **15** | Family houses 4 | 1 789 | grab | No |
| **16** | Apartment buildings 4 | 6 457 | grab | Yes |
| **17** | Apartment buildings 5 | 12 085 | grab | No |
| **18** | Apartment buildings 6 | 3 453 | grab | No |
| **19** | Hospital area | --- | grab | No |
| **20** | City center 2 | 9 738 | grab | No |
| **21** | University dormitories, Apartment houses | 1 874 | grab | Yes |
| **22** | Bus terminal | 1 318 | grab | No |
| **Major Prague's sewer's sampled from 22 March through 21 May 2021** |  |  |  |  |
| **S1** | Main sewer ACK – main influent to Prague's central WWTP | n.a.* | 24-hours composite |  |
| **S2** | Main sewer C | n.a.* | 24-hours composite |  |
| **S3** | Main sewer F | n.a.* | 24-hours composite |  |
| **S4** | Main sewer K | n.a.* | 24-hours composite |  |

### Description of the neighborhoods monitored in this study

The local neighborhoods (Locations 1 to 22) monitored in this study (Fig. 1A) were selected based on their epidemiological importance to achieve an appropriate diversity of wastewater sources. The main sewers (Fig. 1B) were selected to cover a large portion of Prague’s wastewater (to obtain data representative for the whole of Prague) and to observe deviations between data obtained from large wastewater sources. Note that wastewater from Sewer ACK, the main influent to Prague’s WWTP, mainly represents the combination of sewers C and K, and therefore to some extent should give similar results.

**Fig. 1.**
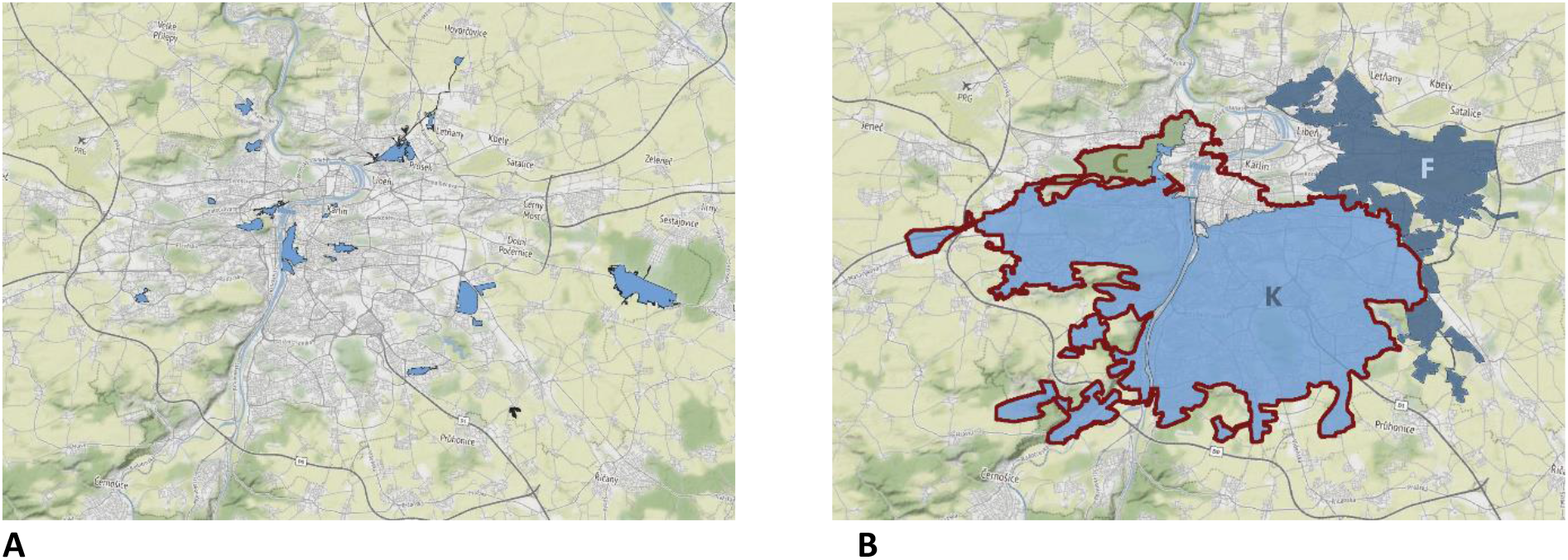
Illustration of the areas served by the sewers sampled in this study: A – Local neighborhoods (the areas on the map are kept anonymous to protect privacy rights); B – Major sewers of Prague ACK, C, F, and K. The red line shows the border of the area served by sewer ACK. This sewer combines sewers A, C, C1, and K and is considered the main influent to Prague’s WWTP.

### Wastewater samples

In local neighborhoods (Locations 1 – 22), 500mL grab samples were taken directly from the sewer and subsequently delivered to the lab within one to two hours. The sampling time was approximately constant for each location and chosen to represent the morning peak (between 7 and 10 a.m.).

From the main sewers (Locations S1 – S4), 1L 24-hour flow-controlled composite samples were taken.

### Samples processing

At the beginning of the monitoring (July through September 2020), the procedure reported by Wu et al. (2020) was used. The sample was first pasteurized at 60 °C for 90 min and subsequently filtered through a 0.45µm nitrocellulose membrane filter (Sigma Aldrich, UK). For each sample, several filters were used due to filter clogging until 40 mL of filtrate was produced. The filtrate was mixed with 4 g of polyethylene glycol (PEG) 8000 (Sigma Aldrich, UK) and 0.9 g NaCl (Penta, Czechia) in a 50mL Falcon tube. After all the contents had dissolved, the tube was centrifuged at 12 000 g and 4 °C for 30 min. The supernatant was discarded and the tubes with precipitate were centrifuged at 12 000 g and 4 °C for another 5 min. The liquid was discarded by pipette, and 500 µL of TRIzol™ Reagent (Invitrogen ThermoFisher, USA) was added to the pellet. The pellet with TRIzol™ Reagent was either directly processed or kept at – 80 °C until RNA extraction (for max. 4 weeks period). After the optimization of the procedure (end of September 2020, data not shown), the pasteurization was omitted and the vacuum filtration was replaced with centrifugation at 4,600 g and 4 °C for 30 min. The supernatant was processed like the filtrate in the procedure above. Another centrifugation step before adding TRIzol™ Reagent, which served for more thorough removal of the rest of the water, was added to the protocol from mid-March 2021. This second centrifugation was performed at 12,000 g, 4 ° C for 5 minutes, the supernatant was removed by pipette, and TRIzol™ Reagent was added to the pellet.

### RNA extraction

After PEG precipitation, TRIzol-chloroform extraction was used for total RNA isolation according to the manufacturer’s protocol (Invitrogen, ThermoFisher Scientific). Since the RNA was finally isolated from a total volume of 80 mL of wastewater, two pellets each initially obtained from 40 mL of wastewater and re-suspended in 500 µL of TRIzol™ Reagent were combined to obtain 1 mL of TRIzol™ suspension. To control the total RNA isolation process, 800 copies of spike ssRNA (EURM-019) were added directly into the sample (suspension of 1 mL of TRIzol™ Reagent and pellets from 80 mL of the sample). Then, 200 µL of chloroform (Penta, Czechia) was added, the sample was vortexed thoroughly for 15 s, and incubated for 13 min at room temperature. After incubation, the mixture was centrifuged at 12,000 g for 15 min at 4 °C in a pre-cooled centrifuge. The aqueous phase containing RNA was transferred into a fresh tube and 500 µl of 2-propanol was added. After 8 min of incubation at room temperature, RNA was pelleted by centrifugation (10 min, 12 000 g, 4 °C). Next, the supernatant was removed and the pellet was washed by adding 1.4 mL of 75% ethanol (Penta, Czechia). The sample was then centrifuged at 7 500 g for 5 min at 4 °C. Finally, the obtained pellet of RNA was briefly dried and resuspended in 50 µL of nuclease-free water (NFW, Top-Bio, Czechia).

### Reverse transcription – multiplex quantitative real-time PCR (RT-mqPCR)

Reverse transcription and PCR amplification were conducted in a single tube at a reaction volume of 20 µl containing a 1x EliZyme OneS Probe Kit (Elisabeth Pharmacon; Czechia), primers (0.4 μM each) and probes (0.2 μM each), nuclease-free water, and 5 µl of RNA in each reaction (undiluted and two-fold dilution). The sequences of all primers and probes used in the multiplex RT-qPCR are listed in Table 2. The oligonucleotides were synthesized by Metabion International AG (Planegg, Germany). Primers and probes used for RT-mqPCR were complementary to the SARS-CoV-2 genome. The sequences for the detection of the N1 nucleocapsid-encoding gene were adopted from the CDC protocol (CDC 2020), for spike protein from the EURM-019 product list (JRC 2020), and for the RNA-dependent RNA polymerase (RdRp) gene from Corman et al. (2020). All RT-mqPCRs were performed using a 7500 Real-Time PCR system (Applied Biosystems, Foster City, California, USA), and the data were analyzed using the 7500 Software v2.0.6. The fluorescence channels were evaluated separately: the FAM fluorophore detected nucleocapsid and spike protein-encoding gene fragments (72 bp and 83 bp, respectively), TAMRA was used for the RdRp of SARS-CoV-2 (100 bp), and HEX was used for the RdRp of SARS-CoV-2, SARS-CoV, and bat-SARS-related coronaviruses (CoVs). Only the FAM channel was used for gene quantification. The serial dilution method was applied for inhibition testing in each sample. Undiluted RNA and 2x diluted RNA (dilution in NFW from Promega, Madison, WI, USA) were used. The samples were analyzed in duplicates. The positive control (target RNA) and no template control (NFW) were run on every RT-mqPCR plate to exclude false (negative or positive) results.

The conditions for transcription and amplifications were as follows: reverse transcription for 10 min at 53 °C, initial denaturation at 95 °C for 2 min followed by 45 cycles of 5 s at 95 °C and 30 s at 60 °C.

The EURM-019 synthetic single-stranded RNA (synthetic ssRNA) fragments of SARS-CoV-2 were used as an *in vitro*-transcribed RNA standard (JRC 2020). This universal synthetic ssRNA of 880 nts contains the target regions that can be amplified by all the RT-qPCR assays listed in Table 1. The procedures for verifying the methodology (quantification, amplification efficiency E, R^2^ coefficient, repeatability, estimation of the Limit of Detection (LOD), and Limit of Quantification (LOQ)) met the criteria of the JRC Technical report on the verification of analytical methods for GMO testing (Hougs et al. 2017). Absolute quantification was carried out by comparing with the standard synthetic ssRNA. Calibration curves were constructed in all experimental runs. The efficiency and R^2^ fulfill the parameters defined in Hougs et al. (2017).

**Table 2:**
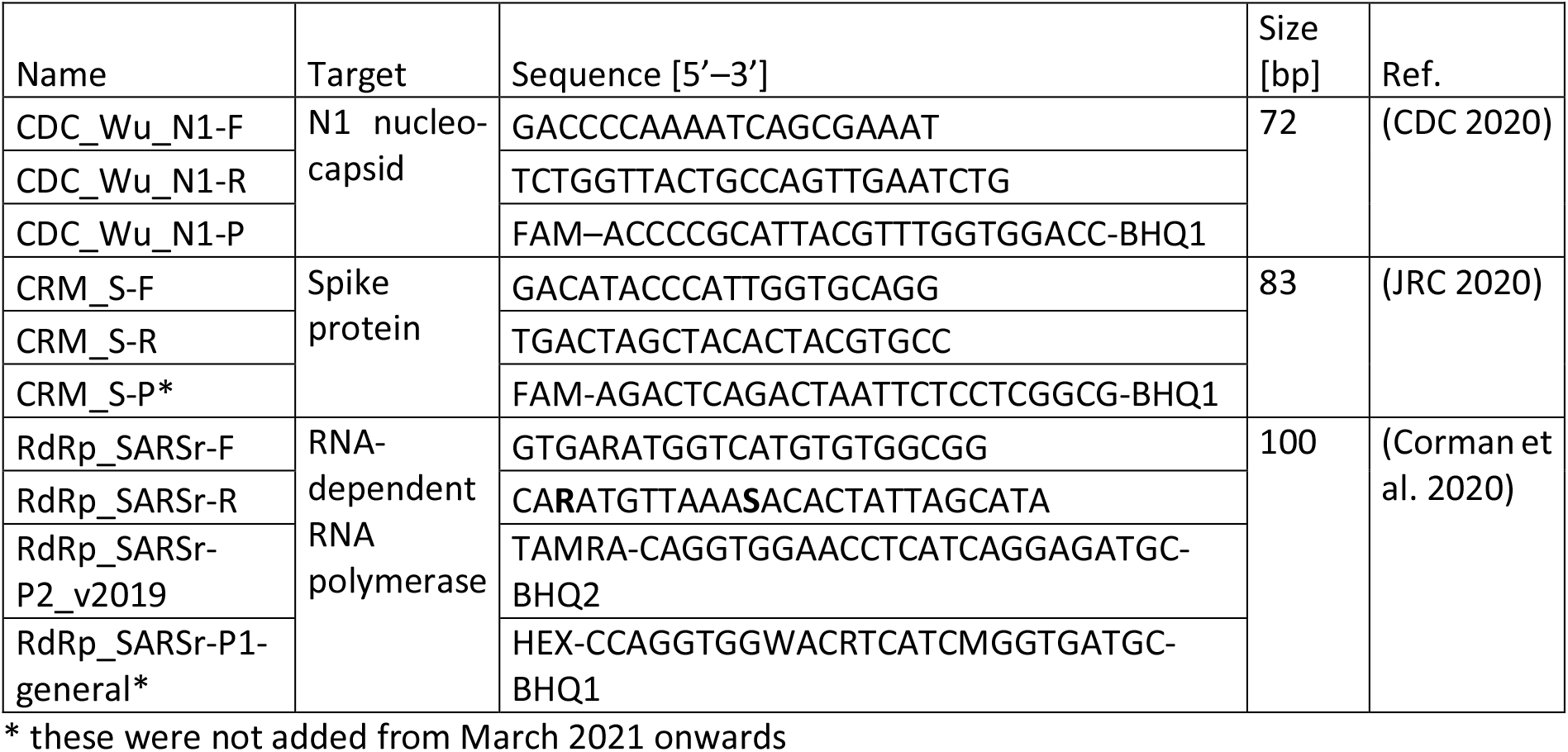
Primers and probes used in this study.

## Results and Discussion

### The course of the COVID-19 epidemic in Prague

After successfully controlling the first wave of the COVID-19 epidemic in spring 2020, the Czech government eased the anti-COVID restrictions during summer 2020. This approach resulted in a massive increase in COVID-positive cases from October to November (a second wave) which was counteracted on 14 October by strict measures that included closing shops, restaurants, and other services, closing most of the schools and student dormitories, restricting free movement and public gatherings, etc. The third wave of new COVID-19 cases in December 2020 and January 2021 followed a partial easing of anti-epidemic measures in December (opening shops). The fourth wave started in February 2021 (probably resulting from the spread of new, more infectious variants of the SARS-CoV-2 virus). The situation in Prague was very similar to the rest of the Czech Republic, and the maximum numbers of newly reported cases per day were as high as 1 708, 2 065, and 2 037 in the second, third, and fourth wave, respectively (Ministry of Health of the Czech Republic 2021). The maximum number of active COVID-19 cases reached 1 506 per hundred thousand inhabitants on 9 March (Fig. 2).

**Fig. 2:**
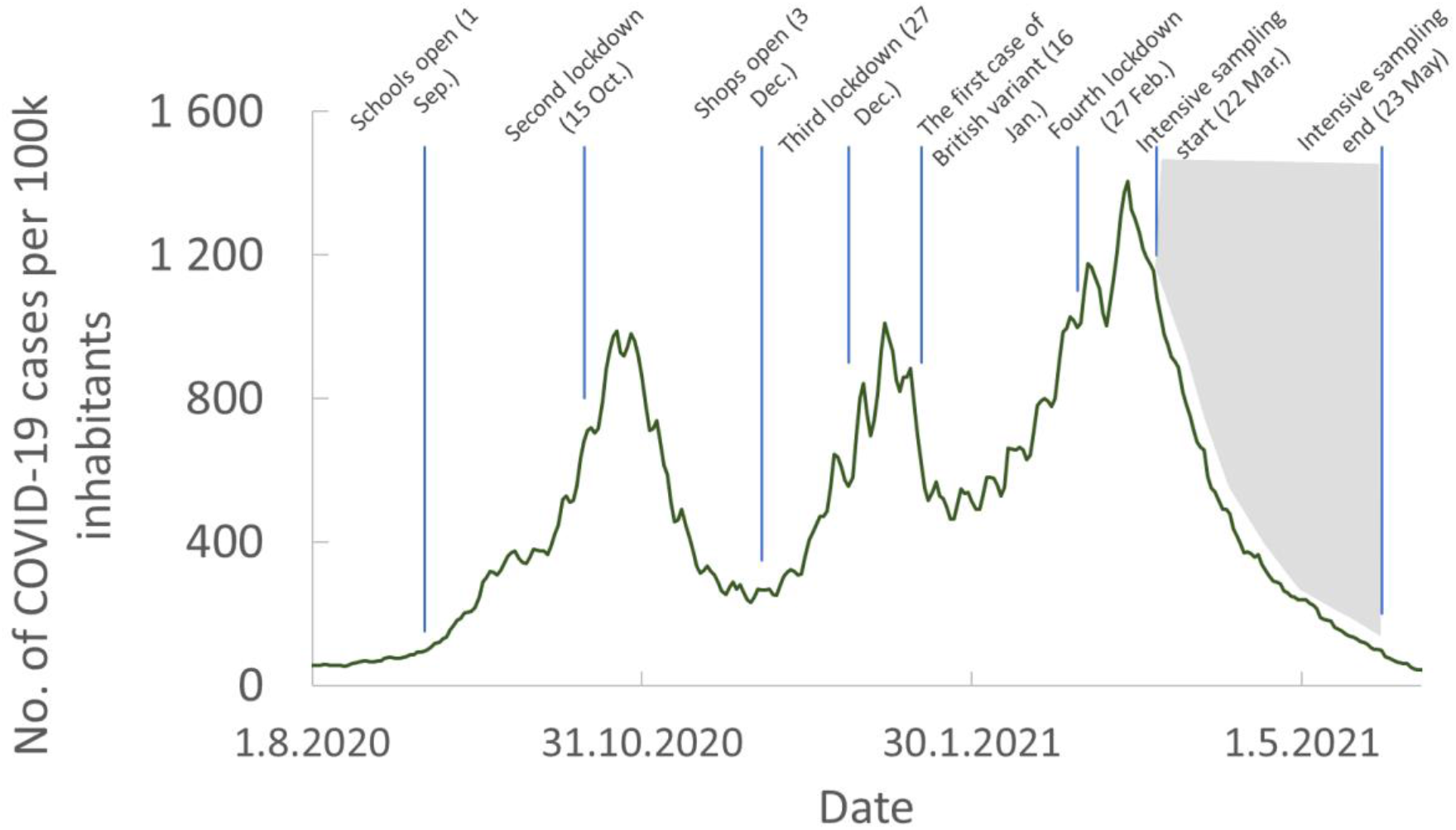
Active cases of COVID-19 disease registered in Prague from August 2020 until the end of May 2021 as reported by the Ministry of Health of the Czech Republic (2021). The blue lines indicate the time of the most important milestones of the epidemic. The grey area marks the period of intensive monitoring.

### Correlation of N1 gene concentration in sewage with epidemic data based on clinical testing

#### The correlation at small sewers in the long-term sampling

The presence of SARS-CoV-2 RNA was detected at all locations of the Prague sewer network chosen for this study. However, the frequency of positive samples, N1 gene concentrations, and the correlation between N1 gene concentration and the number of positive COVID-19 cases were dramatically different between different locations (<u>Fig.</u> 3). Overall, 46% of all tested samples were positive, 47% were negative and 7% could not be evaluated due to PCR inhibition. The relatively large portion of negative samples is given by the fact that a disproportionally high number of the samples were collected during the period of the declining epidemic (22 March – 23 May 2021, Fig. 2.). The numbers of N1 gene copies in the positive samples ranged from 10^1^ to 10^7^ per L of wastewater.

To demonstrate the value of the data measured in wastewater for the monitoring of COVID-19 epidemics, N1 gene copy numbers per L of wastewater on a logarithmic scale were correlated to the estimated active cases in Prague based on clinical testing (Fig. 4). Only the positive samples were taken into account for the correlation analysis.

The highest correlation between N1 concentration in wastewater and clinical epidemic data was observed at locations 3 and 10 (determination factor R^2^ higher than 0.5). As shown in Fig. 4, N1 gene concentrations correlated well (determination factor R^2^ typically from 0.35 – 0.55) with the clinical epidemic data in predominantly residential areas with more than 1 000 inhabitants (Locations 3, 5, 6, or 7) and the residential area dominated by a shopping mall (Location 10, Fig. 4C).

A surprisingly good agreement was observed among the trend lines obtained for all residential areas except for Location 6 (Fig. 4F), showing that the correlation equations can be extrapolated from one location to another. Indeed, the correlation equation obtained for Location 3 was used to estimate the number of active COVID-19 cases per 100 thousand inhabitants for all the other small locations (Location 1 to 22). The good agreement with the data based on clinical testing is demonstrated in Fig. 3 for selected locations. This suggests that wastewater-based data can in some cases (e.g. very small areas or areas with a prevalence of unregistered individuals) be more realistic than the data based on clinical testing, because even if certain persons are identified by clinical testing as infected, localizing their activities is often impossible.

**Fig. 3:**
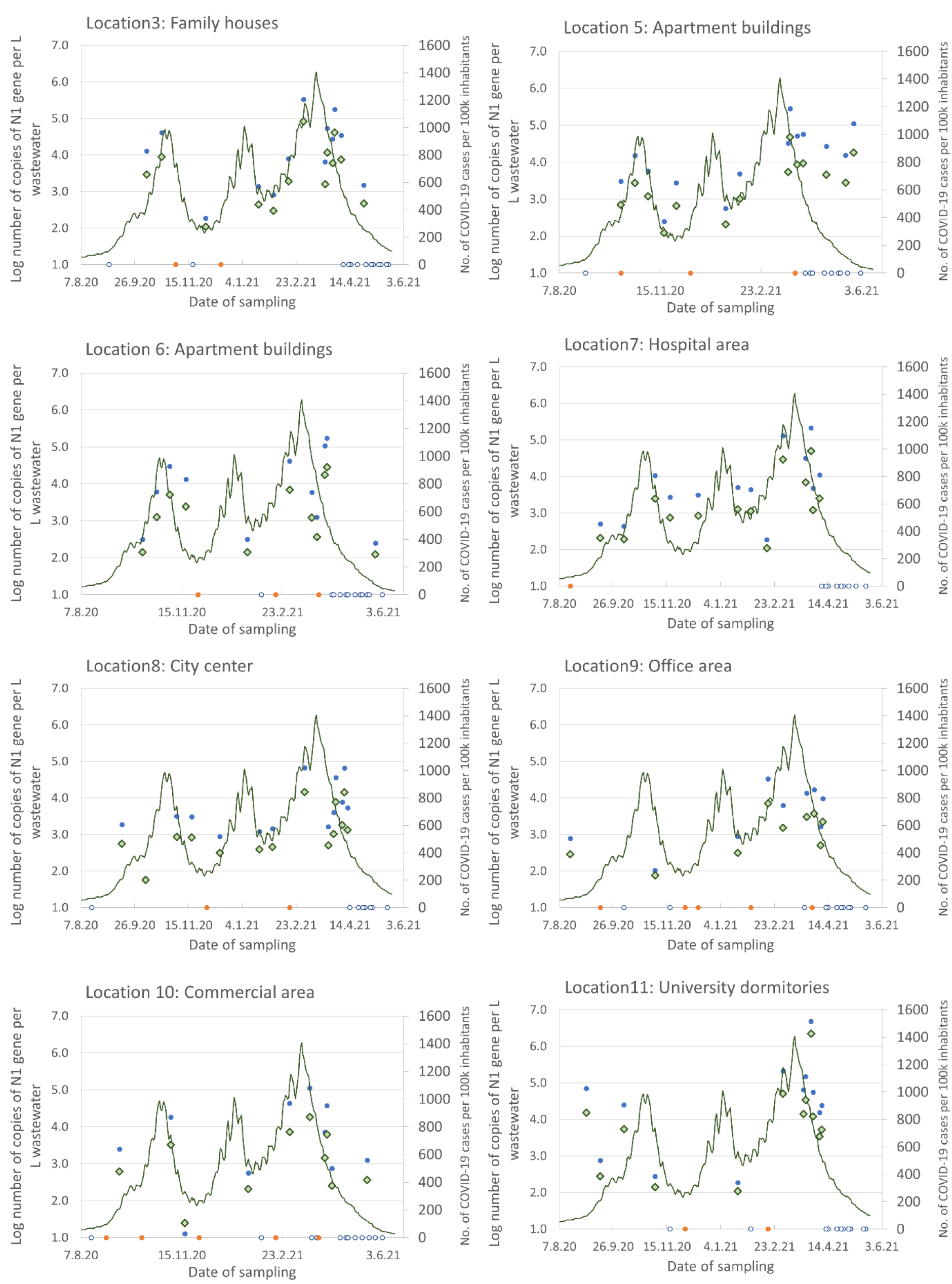
SARS-CoV-2 N1 gene concentration in wastewater (logarithmic scale) and the number of registered positive cases of COVID-19 as reported by the Czech Ministry of health at selected locations monitored since August 2020. A green line – positive cases in Prague per 100 thousand inhabitants estimated from clinical data; Filled blue points – decimal logarithm of N1 gene copy number per liter of wastewater; Empty blue points – negative samples (N1 gene under detection limit); Orange points – samples with unknown PCR inhibitors; Green diamonds – number of positive cases per 100 thousand inhabitants estimated based on N1 gene copy number in wastewater (based on the correlation shown in Fig. 4).

Unlike with large predominantly residential areas, the correlation was weak at most locations dominated by activities other than regular living such as the city center (Location 8, Fig. 4G), student dormitories (Location 11, Fig. 4H), office area (Location 9), or at the airport WWTP (Locations 13 and 14). A weak correlation was also observed in the small residential areas (Locations 1, 2, and 4). E.g. the samples from Location 1 representing a confined luxury residential resort were predominantly negative or with low copy numbers (up to 10^2.3^ copies per L wastewater).

**Fig. 4.**
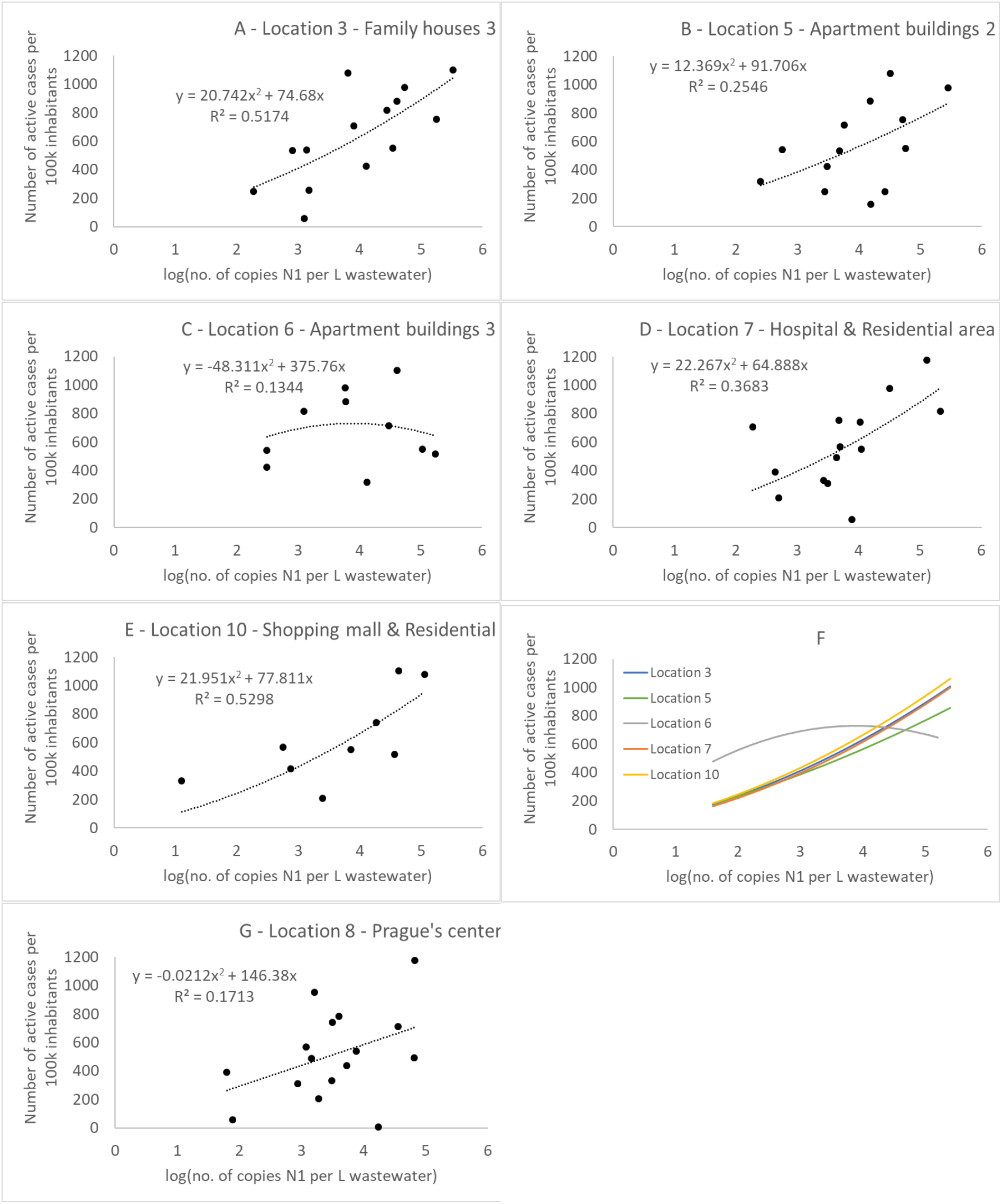
Correlation between N1 gene copy number in wastewater (in logarithmic scale) and the number of active cases of COVID-19 in Prague in small locations where grab samples were collected. A – D: Residential areas with more than 1 000 officially registered inhabitants; E: Area combining shopping mall and residential area; F: The comparison of the polynomic trend lines for larger residential areas (over 1 000 residents) derived from figures A – E. G – An example of a location that is not predominantly residential.

It was not clear whether a close correlation between the copy number of N1 gene detected in wastewater and epidemic data should be expected for all monitored locations. Besides the fact that the real number of infected individuals active at a given location was unknown, the effect of time must also be taken into account when interpreting the data. On some occasions, wastewater-based data exhibit a 1 to 2 weeks lead (Location 11 – student dormitories – in the second wave and Location 3 and 5 – residential areas – in the third wave) or delay in epidemic progression compared to the epidemic data (Location 6 – residential area – in the second wave, Fig. 3).

Some studies highlighted the predictive value of SARS-CoV-2 RNA monitoring in wastewater, claiming that the increase in copy numbers in wastewater (Ahmed et al. 2020, Haramoto et al. 2020, Sherchan et al. 2020, Wu et al. 2020a) or sewage sludge (Peccia et al. 2020) precedes the rise of clinically detected COVID-19 cases by up to two weeks. Medema et al. (2020) detected SARS-CoV-2 RNA in sewage from Amersfoort (the Netherlands) 6 days before the first clinical cases were reported, and La Rosa et al. (2021) showed that the virus was circulating in Italy even in December 2019. Similarly, during the SARS epidemic in 2002, Wang et al. (2005) detected coronavirus RNA originating from the feces of patients hospitalized with SARS-CoV infection in sewage up to 8 days prior to the outbreak of the epidemic.

Unfortunately, this predictive value was not consistently demonstrated in our research. The reason for this observation could be the low frequency of sampling, which was chosen in this study for capacity reasons.

#### The correlation observed in the main Prague sewers and small sewers during the intensive sampling campaign

The major sewers, representing large regions of Prague, were only monitored during the intensive sampling campaign (22 March – 21 May 2021, Fig. 2), when the COVID-19 epidemic had already sharply declined. However, the logarithmic correlation between N1 gene copy numbers and the number of active COVID-19 cases in Prague was as good as for the best-correlating residential areas with a determination factor R^2^ from 0.48 to 0.68 (Fig. 5). Again, the trend lines obtained for individual main sewers were very similar to each other (Fig. 5E), and the calibration performed with the data from Sewer 1 (Fig. 5A) was successfully used to estimate the number of active COVID-19 cases for all the other main sewers (Fig. 5B, C, and D).

**Fig. 5.**
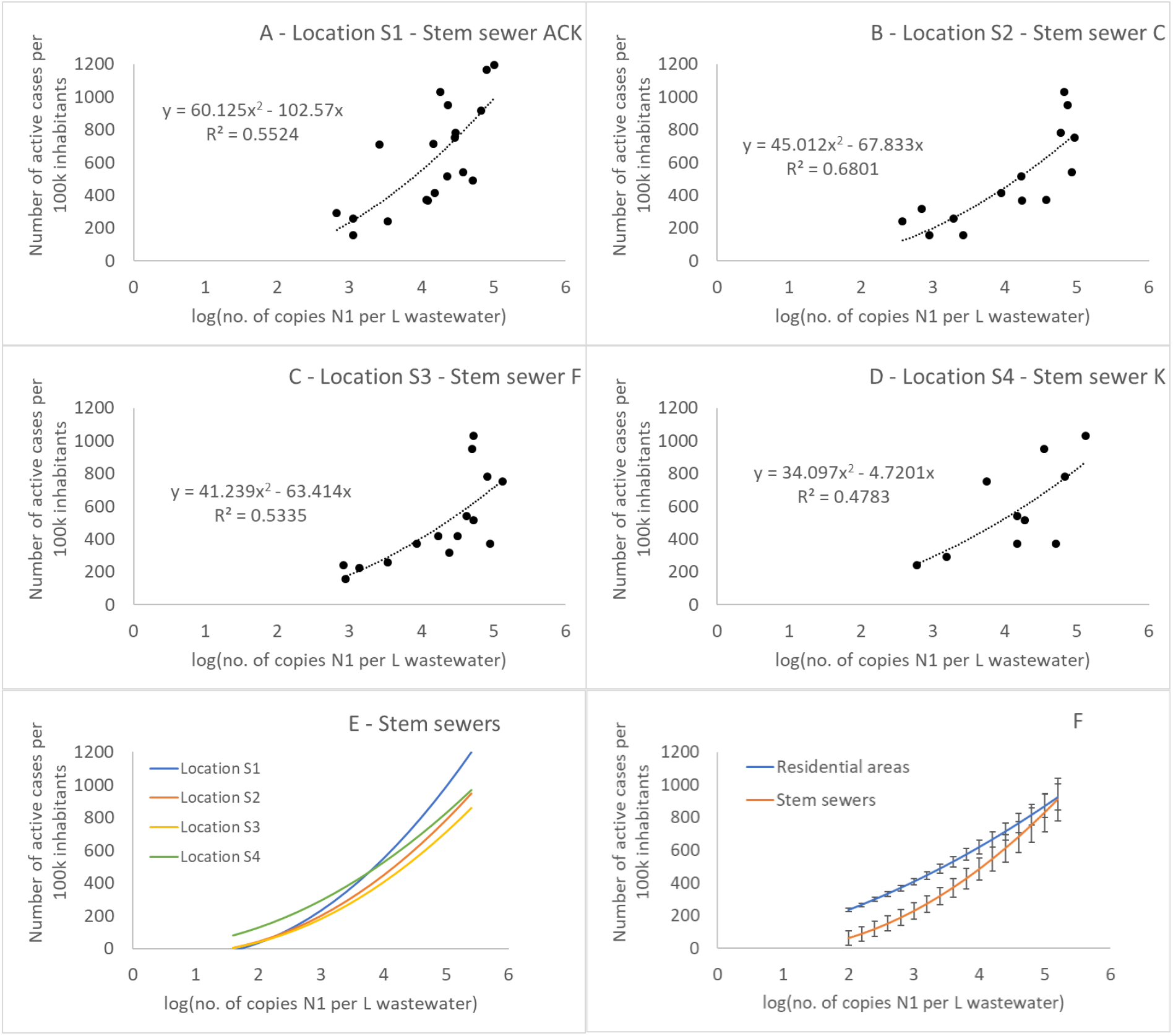
Correlation between N1 gene copy number in wastewater measured in composite 24h samples collected at main Prague sewers and the number of active cases of COVID-19 in Prague. A – D: Main sewers ACK, C, F, and K; E: The comparison of the polynomic trend lines derived from figures A – D; F: The comparison of averaged trend lines for the residential areas (3, 5, 7, and 10) and the main sewers. The error bars represent standard deviations between individual locations.

The averaged trend line obtained for larger residential areas (Locations 3, 5, 7, and 10) was less steep than the trend line for the main sewers (Fig. 5F). Besides other factors, this was given by the fact that at the respective locations that cover in total less than 2.5% of Prague’s population, there was a very low probability of the occurrence of positive cases during the periods with low COVID-19 incidence. Indeed, few positive samples were detected at Locations 1 to 22 when the number of positive cases in Prague was lower than 300 per 100 thousand inhabitants (Fig. 3).

The data for the main sewers came almost exclusively from the period of epidemic decline, when the wastewater-based data were biased by the long excretion of viral RNA in stools, which might persist for up to 2 weeks after the patient is considered COVID-19-negative (Hong et al. 2021, Wu et al. 2020b). The decline in SARS-CoV-2 RNA occurrence in Prague wastewater lagged up to 2 weeks behind the decrease in positive samples in clinical data (Fig. 6E). Nevertheless, positive samples were observed in all main sewers, even though as little as 100 positive cases per 100 thousand inhabitants were registered based on clinical testing (Fig. 6). Interestingly, following the week of 2 – 8 May, when all samples from the main sewers were negative, the numbers of N1 gene copies in the samples from the main sewers started to grow again. Clinical data has not confirmed this trend by the time of writing this report.

**Fig. 6.**
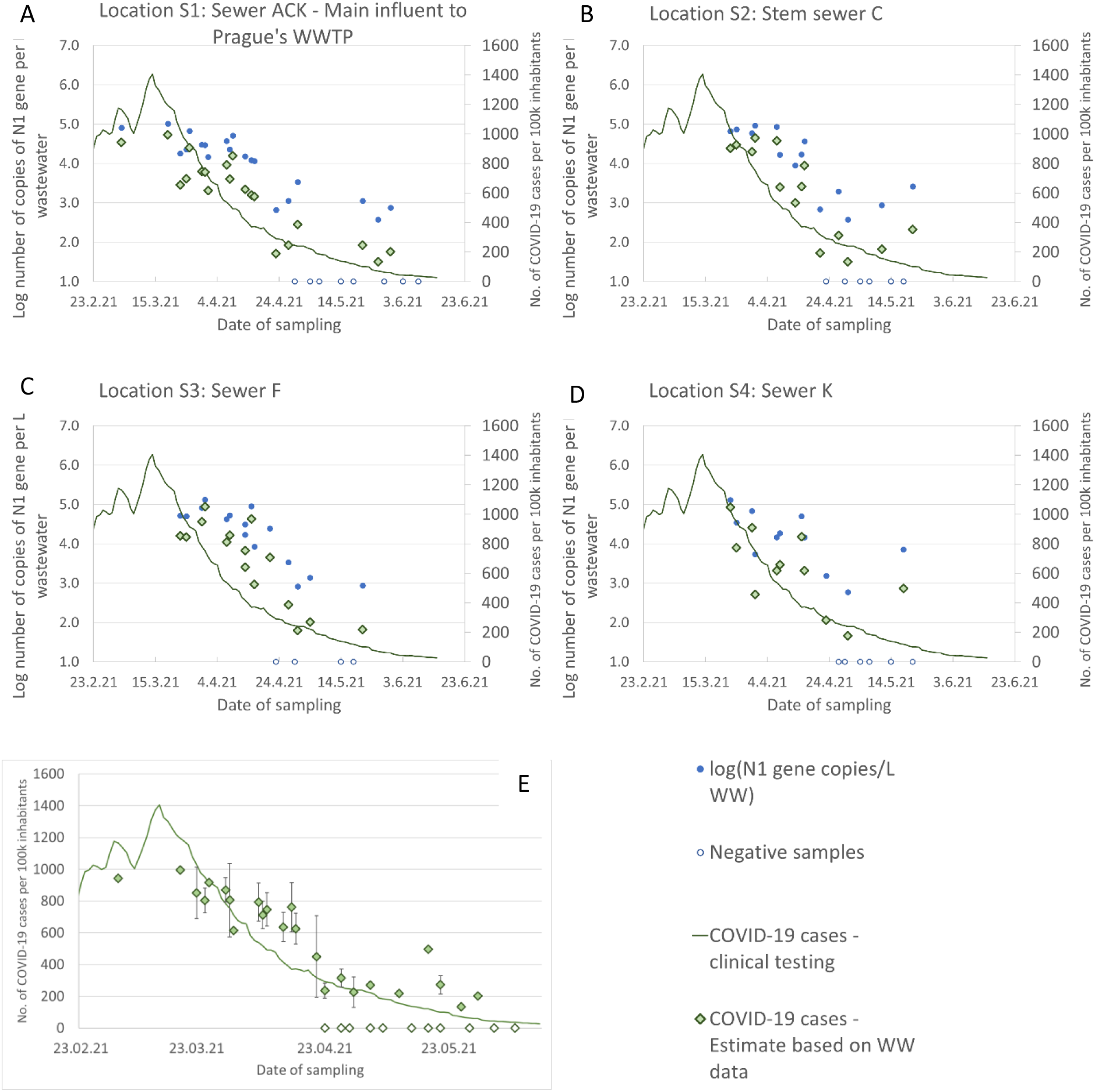
Decline in the presence of SARS-CoV-2 RNA in wastewater during the so-far final phase of the COVID-19 epidemic in Prague, as observed in the major (stem) sewers individually (A – D) and the average from COVID-19 cases estimates for all main sewers (E). Blue dots -number of N1 gene copies per L of wastewater (logarithmic scale); Blue open circles – negative samples; Green diamonds – Number of COVID-19 cases (estimate based on wastewater data); Green line – Number of COVID-19 cases (estimate based on clinical testing).

The decrease in N1 gene copy numbers in the samples collected at small locations (Locations 1 – 22) was generally not delayed compared to the clinical data (Fig. 3, e.g. Locations 3, 7, 8, 9, 10, 11, and others). However, positive samples were repeatedly identified at locations such as 5 (Family houses; Fig. 3), 19 (Hospital; data not shown), and 22 (Bus terminal; data not shown), even while the total number of positive cases in Prague were well below 100 per 100 thousand inhabitants. This observation indicated that problematic areas with the occurrence of infected individuals could be identified despite the low average epidemic numbers. As a result, targeted epidemic measures can be applied in these areas.

### Epidemiological value of wastewater-based data on SARS-CoV-2 occurrence in Prague wastewater

After the very recent paper by Mota et al. (2021) reporting decentralized monitoring of the sewers of Belo Horizonte (Brazil) for three months in 2020, this is the first European study comparing the epidemic relevance of data collected during 10 months at local neighborhoods of different sizes (approx. 100 to 14 000 registered residents) and areas covering large parts (main sewers S2, S3, and S4) or almost an entire city (main sewer S1) of 1.32 mil. inhabitants. The correlation between wastewater-based and clinical testing-based data is surprisingly good for residential areas with more than 1 000 registered inhabitants and is approximately as good as the same correlation obtained for the main sewers. Moreover, the data obtained from the small residential areas seemed to be quicker than those from the major sewers, at least during the decline of the COVID-19 pandemic.

Mota et al. (2021) showed that COVID-19 hotspots could be identified in the city based on data generated by decentralized sewage monitoring especially in vulnerable neighborhoods such as favelas, which tend to be densely populated and with limited sanitation infrastructure. Our study shows on long-term data that epidemic hotspots escaping the attention of governmental authorities may occur also in a major European city where such vulnerable areas are not expected.

Huisman et al. (2021) showed the feasibility of Reproduction factor (R_e_) estimation from the analysis of SARS-CoV-2 RNA in wastewater independently of clinical data. The correlation between the results of wastewater analyses and epidemic data in Prague published in this article supports this conclusion. This finding is very important, as the estimation of R_e_ from wastewater can be done in real time, is robust, rapid, low cost, and applicable for various wastewater matrices.

Last, but not least, we showed that grab samples could be as informative as 24h composite samples. This might be due to precise time-dependent sampling (typically during the morning peak) and avoidance of the effects of weather conditions (the samples were not collected shortly after major precipitation events). As the correlation between the number of N1 copies and COVID-19 cases was logarithmic, regular variations in wastewater flow (even hundreds of %) had a negligible effect on the numbers of gene copies per L of wastewater. Indeed, also Black et al. (2021) showed that grab samples may be of high epidemiological value when the time of sampling is chosen approprietly.

The representativeness of the concentration-based data was demonstrated by a comparison of the correlation for gene N1 copies per L of wastewater and gene N1 copies per hour (concentration multiplied by average daily wastewater flow; Fig. 7), which is often used for correlation with clinical-based data (Huisman et al. 2021). The determination factor (R^2^ = 0.481) was almost identical to the one obtained for the concentration of N1 gene copies (R^2^ = 0.478), shown in Fig. 5D. The same was true for the other main sewers (data not shown). Therefore, the measurements of SARS-CoV-2 RNA concentration seem to be sufficient for the estimation of COVID-19 cases, even without knowing the exact wastewater flow at the time of sampling. Additionally, these data are usable for epidemic monitoring without using complex mathematical models such as the one presented by Hart and Halden (2020).

**Fig. 7.**
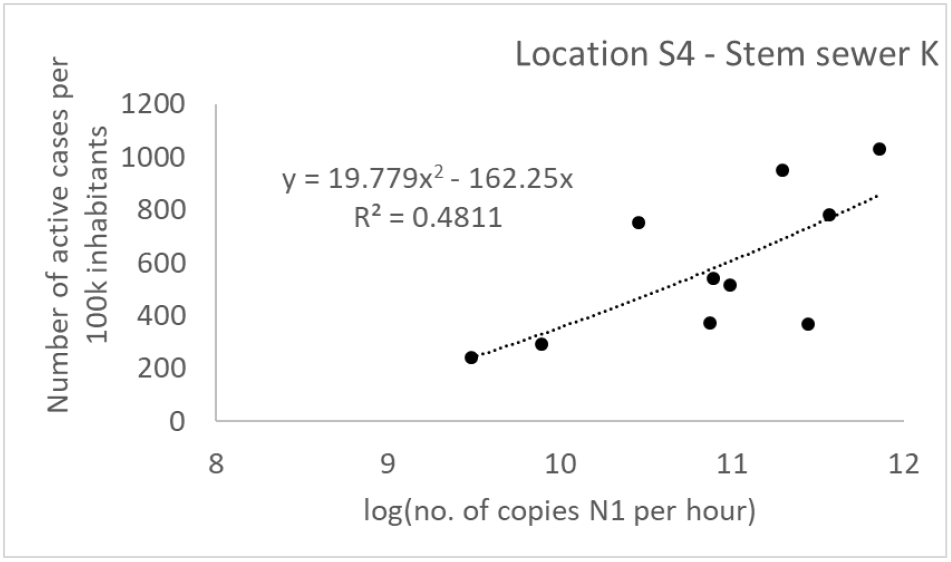
Correlation between N1 gene copy number detected per hour in wastewater sampled from one of the main Prague sewers and the number of active cases of COVID-19 in Prague.

### The inhibition of RT-qPCR

Wastewater is a complex matrix that contains several potential inhibitors of reverse transcription as well as PCR. Such inhibitors include humic acids, fulvic acids, humic material, metal ions, polyphenols from the environment, and complex polysaccharides, bile salts, lipids, and urate from stools (Schrader et al., 2012). These inhibitors of the molecular biology assay should be removed during sample preparation for RT and PCR, mainly during RNA isolation.

We used the TRIzol™ isolation method, as it is reliable and relatively inexpensive. Other studies also recommended this method for municipal wastewater samples. E.g. Torii et al. (2021) observed the highest recovery of phage RNA while using pre-concentration by PEG followed by TRIzol™ RNA isolation. Even though the RT-qPCR was inhibited in some samples collected in our study, this was relatively rare and did not jeopardize the interpretation of our data.

## Conclusions

This paper reports the presence of SARS-CoV-2 RNA in wastewater collected at different locations within a large city, such as residential areas of various sizes, hospitals, city center areas, student dormitories, transportation hubs, and commercial areas within a large city (Prague, Czechia). The data obtained at the main Prague sewers were more consistent than those obtained from the small sewers. However, the correlation between wastewater-based data and those from clinical testing was good for the residential areas with more than 1 000 registered inhabitants. The latter was true despite the fact that only grab samples were collected from the small sewers. This study also shows that monitoring SARS-CoV-2 RNA in wastewater sampled from small sewers can identify an isolated occurrence of COVID-19-positive cases in local neighborhoods. This can be highly valuable while tracking COVID-19 hotspots within a large city.

## Data Availability

not relevant

## Acknowledgments

This work was financially supported by Czech Technological Agency TA CR grant no. SS01020112 “Technology for the removal of antibiotic resistance from sewage sludge applied in agriculture”. The authors also wish to thank PVK a.s., VZU, and UCT Prague for their generous financial and material support. We also wish to thank Jiri Jarkovsky of the Institute of Health Information and Statistics of the Czech Republic for providing epidemic data for the areas under study. The valuable assistance of Karel Behounek (Aqua-Contact, v.o.s.) and Irena Novakova (Prague Airport) during wastewater sampling is greatly appreciated, as well as the dedicated lab work of Nelly Matouskova (Czech Health Institute), Marco Lopez, Sara Doubovska, Katarina Hanusova, and Dominik Tomanek (students of UCT Prague). Finally, we thank to Christof Uisk (student of UCT Prague) for creating the maps of the catchment areas of all sewers.

